# Normal Childhood Brain Growth and a Universal Sex and Anthropomorphic Relationship to Cerebrospinal Fluid

**DOI:** 10.1101/2020.05.19.20102319

**Authors:** Mallory R. Peterson, Venkateswararao Cherukuri, Joseph N. Paulson, Paddy Ssentongo, Abhaya V. Kulkarni, Benjamin C. Warf, Vishal Monga, Steven J. Schiff

## Abstract

**Object:** The study of brain size and growth has a long and contentious history, yet normal brain volume development has yet to be fully described. In particular, the normal brain growth and cerebrospinal fluid (CSF) accumulation relationship is critical to characterize because it is impacted in numerous conditions of early childhood where brain growth and fluid accumulation are affected such as infection, hemorrhage, hydrocephalus, and a broad range of congenital disorders. This study aims to describe normal brain volume growth, particularly in the setting of cerebrospinal fluid accumulation.

**Methods:** We analyzed 1067 magnetic resonance imaging (MRI) scans from 505 healthy pediatric subjects from birth to age 18 to quantify component and regional brain volumes. The volume trajectories were compared between the sexes and hemispheres using Smoothing Spline ANOVA. Population growth curves were developed using Generalized Additive Models for Location, Scale, and Shape.

**Results:** Brain volume peaked at 10-12 years of age. Males exhibited larger age-adjusted total brain volumes than females, and body size normalization procedures did not eliminate this difference. The ratio of brain to CSF volume, however, revealed a universal age-dependent relationship independent of sex or body size.

**Conclusions:** These findings enable the application of normative growth curves in managing a broad range of childhood disease where cognitive development, brain growth, and fluid accumulation are interrelated.

## Introduction

The brain growth and CSF accumulation relationship is critical to characterize because it is impacted in numerous conditions of early childhood where brain growth and fluid accumulation are affected such as infection, hemorrhage, hydrocephalus, and a broad range of congenital disorders ^1,2^. Appropriate management of such diseases requires understanding the normal dynamics of brain growth in relationship to fluid accumulation. However, the study of brain size and growth has a long and contentious history^3,4^. In the pre-magnetic resonance imaging (MRI) era, post-mortem studies provided insight into brain volume changes over the lifespan, but such methods suffered from inherent inaccuracies^5^. In the MRI era, the development of advanced computational and statistical algorithms has enabled detailed in-vivo volumetrics including components and regions of the brain^6-9^.

Studies defining normal brain volume growth patterns in the MRI era have often included small sample sizes, limited algorithm technology, incomplete coverage of the pediatric age range, retrospective cohorts taken from clinical patients, and a disparate array of curve fitting techniques^6-8,10,11^. Studies conducted by Pfefferbaum et al (1994), Giedd et al (1999), and Courchesne et al (2000) included only 30, 243, and 50 subjects in the pediatric age range^6,7,9^. Secondly, none of those studies included the entire pediatric age range from birth to age 18. More recent works such as Coupe et al (2017) and Hedman et al (2012) have incorporated big data or meta-analysis approaches, but by incorporating such large amounts of data, they lose rigor in exclusion criteria, population representation, and calibration between scan site acquisition^12,13^. Furthermore, despite leveraging 56 studies and including 2,211 participants, Hedman et al (2012) did not include any subjects under the age of 4, excluding the period during which the majority of brain growth takes place^12^. Others such as Knickmeyer et al (2008) have looked closely at the infantile period from birth to age 2, but have not covered the later pediatric ages^14^.

Despite this enormous amount of recent interest, a definitive set of percentiled normative curves for brain growth are not yet available as we have for multiple other anthropomorphic measures such as weight, height, and head circumference. Furthermore, the interaction of brain growth and the regulation of the fluid compartment within which it is buoyant and immersed during development has not been well characterized. We here resolve both of these open issues.

## Materials and Methods

### Cohort Characteristics

The MRI scans used in this study were taken from the NIH Pediatric MRI Repository (https://nda.nih.gov) under an institutional data use agreement (214908) between The Pennsylvania State University and the National Institute of Mental Health approved on January 14, 2019. This repository was developing using a scaled down United States (US) census from 2000 (in order to appropriately represent the racial and socioeconomic demographic characteristics of the entire US pediatric population) and included rigorous exclusion criteria to ensure healthy participants with normal brain development. The repository aimed to achieve two-year longitudinal follow-up scans for individual participants, and was able to accomplish this for 378 of the subjects, making this a cross-sequential study^15^. The cross-sequential format is ideal for the development of growth curves^7,16^. The number of subjects in this study was 505 (259 female), with a total of 1067 MRI scans due to the longitudinal nature of the cohort. The minimum age was 13 days, and the maximum age was 22 years, but only scans from subjects up to 18-years-old were included to develop growth curves representative of the pediatric age range. Scans existed for participants in each year of life throughout the pediatric age range, as seen in Figure 1.

**Figure 1.**
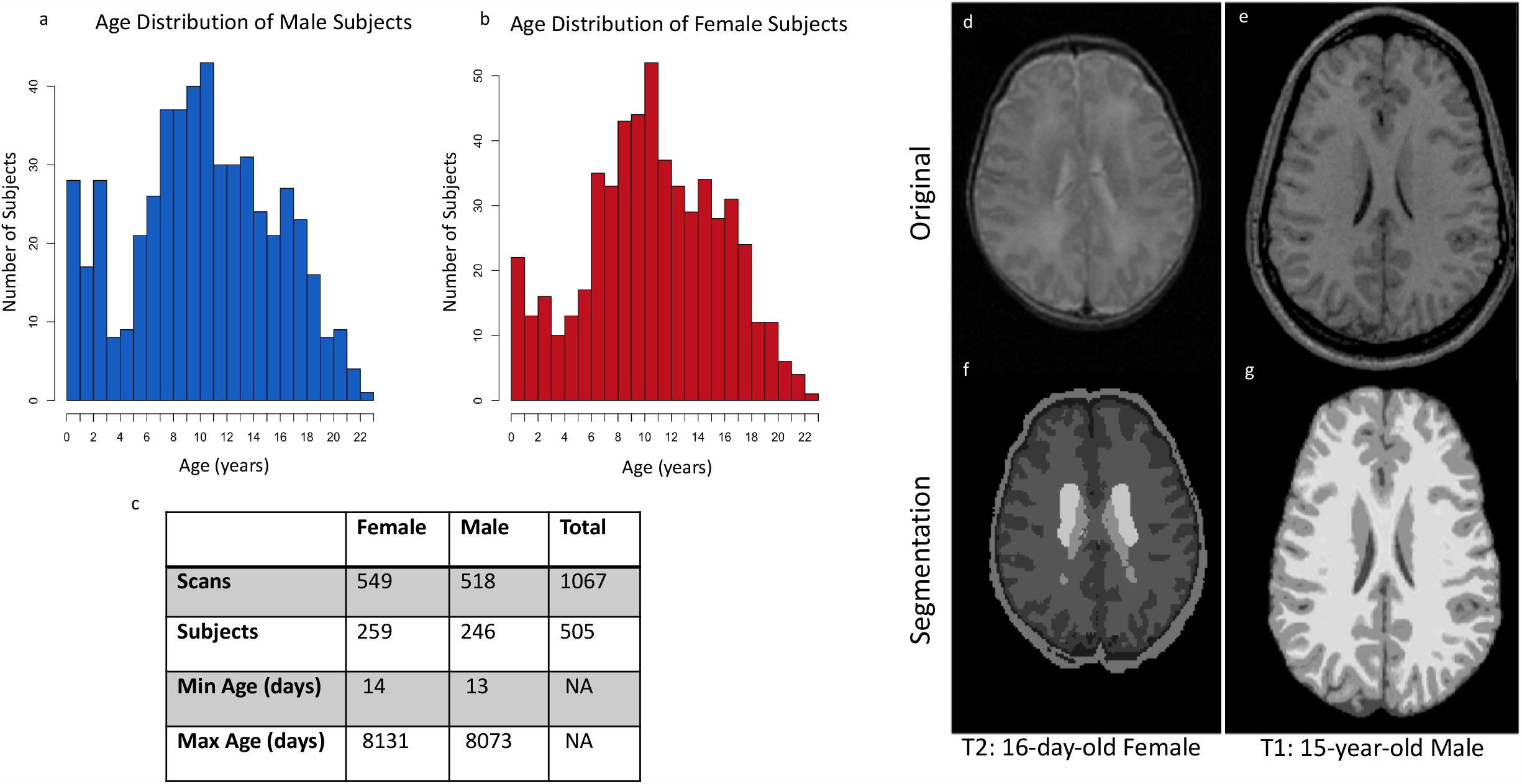
Cohort Characteristics. Data from each age group from 13 days to 22 years was included in the cohort, for both **a)** males and **b)** females. The cross-sequential study included **c)** 505 total subjects, with most subjects having between 2 and 3 longitudinal visits (separated by 2 years). For this study, only scans from 13 days to 18 years were included in order to focus on the pediatric age range. Two different algorithms for volume quantification were used, one for the neonates and one for the older cohort. As an example, a **d)** T2 MRI scan from a 16-day-old female is shown. This scan was processed through the dHCP pipeline to produce a **f)** segmentation including grey matter (found by adding the deep grey matter and cortical grey matter), CSF, and white matter. On the right, the **e)** T1 MRI scan from an older patient (a 15-year-old male) is shown. This scan was processed using the CAT12 pipeline within SPM to produce the **g)** segmentation including grey matter, CSF, and white matter.

### MRI Acquisition and Quality Control

The Pediatric MRI Repository database was collected at Boston Children’s Hospital, Cincinnati Children’s Hospital Medical Center, University of Texas Houston Medical School, the Neuropsychiatric Institute and Hospital UCLA, Children’s Hospital of Philadelphia, and Washington University in St. Louis. While it is of benefit to use multiple different geographic locations when cultivating a cohort representative of a particular population that can then be leveraged for growth curves, bias can be introduced through the use of different scanner sites.

MRI conditions such as scanner manufacturer, acquisition protocols, and field strength can all contribute to the underlying compatibility of MRI data gathered at different sites. For the Pediatric MRI Repository, the acquisition protocols were well-defined and reproduced uniformly at 1.5T at each separate site^15^. Additional acquisition details can be found in the Supplementary Material.

Because different MRI scanners were used the same healthy person was scanned at each site to serve as a calibrating ‘living phantom’^17^. In order to ameliorate the inter-site differences, and characterize the gradient distortion field, the investigators applied a geometric distortion correction using a 3D American College of Radiology (ACR) phantom. The ACR phantom scanning was completed approximately monthly and the living subject scanning was completed annually to ensure inter-site calibration.

In regard to the quality control of the dataset, each MRI scan within the Repository was viewed by a radiologist to ensure no neurologic abnormalities, and the scans were then assessed for quality (in addition, if the first scan taken had too much motion artifact it was redone). If the scan was deemed to be neurologically normal and of acceptable quality, only then was it included in the database.

In our present work with this database, all 1381 available anatomic MRI scans from the database were analyzed. Each volume segmentation was then manually audited to ensure reliable results. Only if each segmentation produced by the pipelines was deemed acceptable upon inspection was a processed scan included in our study. We found 1067 of the 1381 scan sets to be successfully processed, leading to our final segmentations used in our analyses.

### Segmentation Algorithms

Currently, no single algorithm exists that can reliably segment both young infants and older subjects^18,19^. This is particularly due to the myelination changes that do not resolve until approximately two years of age^20^. These myelination changes lead to difficulty in establishing intensity thresholds between grey matter, white matter, and CSF^20^. In order to maximize thresholding intensities, neonatal segmentation techniques most commonly rely on T2 weighted MRI scans, rather than T1 weighted scans that are the predominant scan type used in older cohort segmentation algorithms. Due to these reasons, two different algorithms were used in this study.

The neonates and infants were assessed using the Developing Human Connectome Project (dHCP) pipeline, which required T2 images. The dHCP pipeline is freely available (https://github.com/BioMedIA/dhcp-structural-pipeline).

The older subjects were assessed using the Computational Anatomy Toolbox 12 (CAT12) within the Statistical Parametric Mapping (SPM) platform using Matlab 2019b, which relies on T1 images^21^. Each of the resulting scan sets was manually curated to ensure that appropriate skull-stripping and segmentation was accomplished. Upon establishing the volumes determined by each segmentation procedure, the accompanying atlases were used to compile volumes for the desired regions from smaller sections of the brain.

### Statistical Analysis: Smoothing Spline ANOVA

One of the major limitations of the historical fitting efforts is the inability to statistically assess which time points throughout the entire growth process are truly different between groups while also establishing appropriate representative trajectories. Within this study, we split our efforts, using one approach for trajectory modeling and a different approach for statistical analysis. The Generalized Additive Models for Location, Scale, and Shape (GAMLSS) approach was used to provide smooth trajectory growth curves and centiles for reference in the same manner that the World Health Organization produces their growth curves. These curves, however, do not facilitate an in-depth statistical analysis of differences between groups such as the two different sexes or hemispheres. Therefore, the Smoothing Spline ANOVA technique was also incorporated into this study for the main purpose of examining longitudinal statistical differences between populations. The smoothing parameters are determined through an iterative cross-validation process, which is then followed by the minimization of the penalized least square functional, the solution of which is a smoothing spline. The details of this process are described in Gu (2014)^22^, and are more fully described in the Supplementary Materials.

For this study, the component differences between males and females were explored within the R platform using non-parametric Smoothing Spline ANOVA mixed-effects models with a subject-specific random effects component added to account for the cross-sequential aspect of the data^22^. The model included age and sex or hemisphere as main factors, as well as an interaction term. We were interested in visualizing and statistically analyzing the longitudinal differences between hemispheres, and therefore incorporated hemisphere as a main factor rather than a nested factor. Time periods with significant sex or hemispheric differences were defined as regions where there was no overlap between the Bayesian 95% confidence intervals calculated for the sex and hemispheric factors. These regions of significant difference were highlighted on the plotted models, and the time period of significance was documented as well.

### Statistical Analysis: Generalized Additive Models for Location, Scale, and Shape (GAMLSS)

The smooth growth curves used to fit the volumes and other growth metrics included in this study were developed using the GAMLSS software implemented in R^23^. The Box-Cox power exponential distribution, which was chosen by the World Health Organization (WHO) for their standard growth curves, was used to model the volumes in this study^24^. For the total brain tissue growth curves, we utilized data for subjects between 18-22 years of age to set the 18 year old volume intercepts, and the perinatal volume data from Huppi et al to set the volume intercepts at birth^25^.

In order to determine the peaks of the brain tissue and grey matter curves, cftool within Matlab 2019b was used to fit differentiable rational polynomial functions (which were applied as the smoothing function in the GAMLSS curves). The significant sex and hemispheric differences were found using the Mann-Whitney U-test within Matlab 2019b. A Bonferroni correction was applied so that significance was established with p<0.000066.

The weight for height and height for age normalizations were accomplished by fitting a GAMLSS curve to the weight for height and height for age data from the NIH repository for each sex. Based on these fits, percentiles were calculated for each subject, and the 50^th^ percentile was set at 1, with percentiles above and below ranging from 0.5 to 1.5. The corresponding brain volume for each subject was then divided by this percentile value to achieve normalized brain volumes.

### Statistical Analysis: Cognitive Score Correlations

Cognitive scoring was included in the NIH Pediatric MRI Repository study, with Wechsler Abbreviated Scale of Intelligence (WASI) Tests undertaken on participants ranging from 6 years of age to 18 years of age. The infants (from birth to 3 years of age) were assessed using the Bayley Scales of Infant Development, Second Edition (BSID-II) Mental Development Index (MDI). We fit a linear mixed effects model (with subject identification as the random effects component) to the appropriate cognitive score using the brain volume z-score values.

While the linear fits showed positive slopes for each metric, only the WASI scores showed a significant fixed effect for the brain volume z-score. Age-dependent correlations were calculated with a subpopulation window-size of 120 and step-size of 20 subjects for raw brain volume, brain volume z-score, and weight-for-height normalized volume z-score.

## Results

We quantified brain and CSF volumes from 1067 MRI brain scans from 505 healthy normal subjects (259 female) from birth to age 18. These MRI scans were collected from healthy pediatric participants in the NIH Pediatric MRI Repository (https://nda.nih.gov), which was accessed under an institutional data use agreement between The Pennsylvania State University and the National Institute of Mental Health. This MRI repository was developed based on a scaled-down US census with rigorous exclusion criteria, with the goal of providing a standard representation of the socioeconomic, male and female, and racial distribution of healthy normal US children^15^. The cross-sequential cohort contains participants in each year of life ranging from 13-days to 22-years old (Figure 1a-c). Uniform MRI scanning protocols were applied at each data collection site, and living phantom calibration was leveraged to ensure volumetric consistency between sites^15,17^.

The T2 MRI images of the neonatal and infant population were processed using the Developing Human Connectome Project (dHCP) Pipeline in order to appropriately segment the rapidly growing and incompletely myelinated brains^26^. The T1 images of the older subjects were processed using the Computational Anatomy Toolbox (CAT) within the Statistical Parametric Mapping (SPM12) software. This software utilizes voxel and region-based morphometry upon application of an adaptive probability region-growing (APRG) algorithm for skull-stripping^21^.

Each of the resulting segmentation images (Figure 1d-g) were manually inspected to confirm appropriate anatomic assignment.

The brain components and region volumes quantified by the dHCP and CAT12 pipelines were then fit using Smoothing Spline ANOVA (SSANOVA) with random effects (to account for the cross-sequential design of the study) to define time periods of statistically significant differences stratified by sex and hemisphere (Figure 2, 3, and Supplemental Figure 1)^22^. In order to define the growth trajectories and population centiles, the volumes were also fit using Generalized Additive Models for Location, Scale, and Shape (GAMLSS) with a Box-Cox power exponential (BCPE) distribution (Supplemental Figures 2 and 3), which is the same platform and distribution leveraged by the World Health Organization to develop their standard growth curves for weight, height, and head circumference^23,27^. Finally, the relationship between cognitive scores and brain volume was explored using a linear mixed effects model and age-dependent correlations (Figure 4 and Supplemental Figure 4).

**Figure 2.**
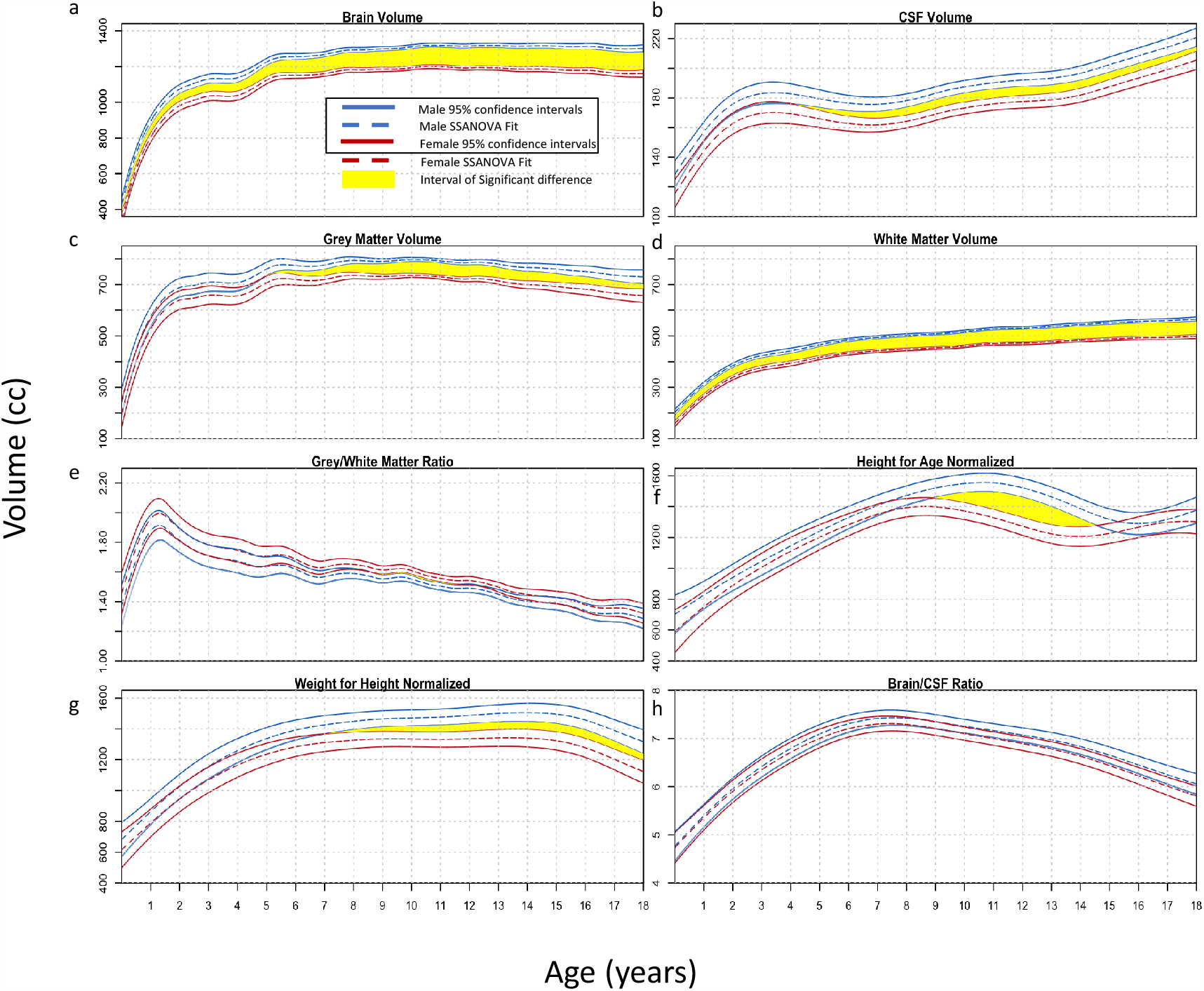
Brain Component Sex Differences. SSANOVA models with random effects were fit to **a)** total brain volume (cc), **b)** cerebrospinal fluid (CSF) (cc), **c)** grey matter (cc), **d)**white matter (cc), **e)** grey to white matter ratio, **f)** height for age normalized brain volume, **g)** weight for height normalized brain volume, and **h)** brain to CSF ratio. For each plot, the male data is shown in blue and the female data in red. The dashed line represents the fit, with the two solid lines on either side showing the Bayesian 95% confidence intervals. The upper and lower intervals for the right and left sides overlap throughout the entire time frame for the brain/CSF ratio, and therefore there are no statistically significant differences at any age. For the other plots, time periods of significant differences where the intervals do not overlap are shown in yellow.

**Figure 3.**
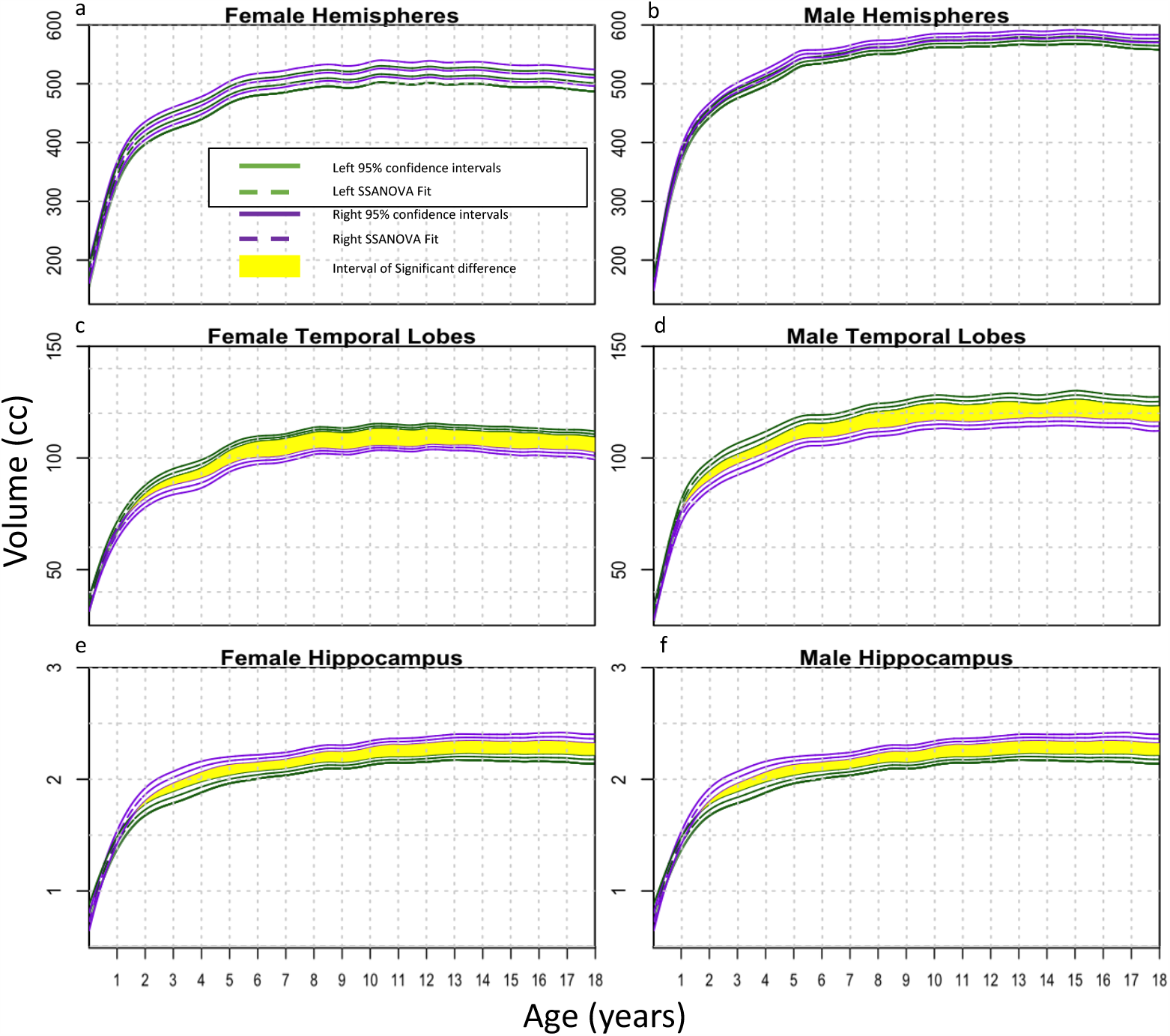
Brain Region Hemispheric Differences. SSANOVA models with random effects were fit to **a**,**b)** hemispheres (cc), **c**,**d)** temporal lobes (cc), and **e**,**f)** hippocampi (cc). Female regional growth curves from birth to 18-years-old are shown in the left column, and the corresponding male regional curves are shown in the right column. For each plot, the left side is shown in green and the right side in purple. The dashed line represents the fit, with the two solid lines on either side showing the Bayesian 95% confidence intervals. The upper and lower intervals for the right and left sides overlap throughout the entire time frame for the hemispheres, and therefore there are no statistically significant differences. For the temporal lobes and hemispheres, time periods of significant differences where the intervals do not overlap are shown in yellow.

**Figure 4.**
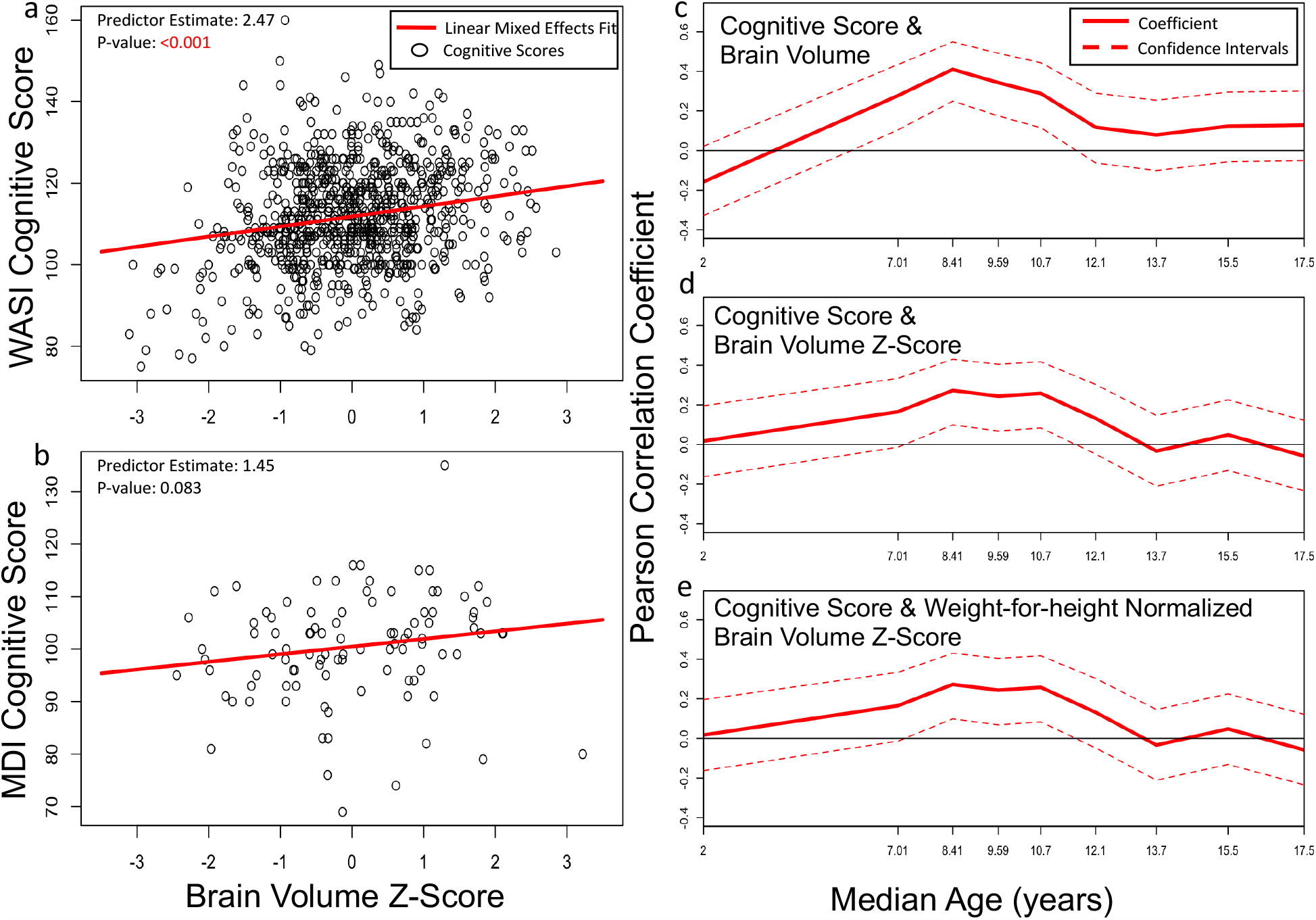
Cognitive Score Correlations. Two cognitive scores were used, the Mental Development Index (MDI) for infants from birth to age 3, and the Wechsler Abbreviated Scale of Intelligence (WASI) Tests for adolescents aged 6 to 18. Linear mixed effects models were fit for **a)** the relationship between MDI and brain volume z-score, as well as **b)** the relationship between WASI and brain volume z-score. Age-dependent correlations were calculated with a subpopulation window-size of 120 and step-size of 20 subjects to show the Pearson Correlation between cognitive score and **c)** raw brain volume, **d)** brain volume z-score, and **e)** weight-for-height normalized brain volume z-score at different ages.

### Findings Differentiated by Sex

Males exhibited larger overall brain volumes than females throughout childhood (Figure 2a). The volume for females peaked at 10.7 years, and the volume for males peaked at 11.2 years, followed by a slow, but consistent decrease. This early adolescent peak has been previously described^10,12^. CSF increased throughout childhood, with male fluid accumulation significantly larger than female after the third year of life (Figure 2b). Grey matter (Figure 2c) peaked at 7.5 years for males and 7.4 years for females and then began to gradually decrease, while white matter (Figure 2d) continued to progressively increase into early adulthood^28^. The ratio of grey/white matter (Figure 2e) showed an initial increase, peaking before 2 years of age and followed by a progressive decrease thereafter. Female grey/white matter ratios were significantly larger than male ratios between ages 9 and 11 although the difference was small (Figure 2e).

We performed body size normalization to assess if differences in brain volume persisted between males and females. Normalizing the brain by body size is not a new concept; allometry, or differential growth, of the brain with respect to body size was discussed in depth by D’Arcy Wentworth Thompson in 1917 in *On Growth and Form*^*29*^. Gould, in *The Mismeasure of Man*, attempted and failed to eliminate sex differences through body size normalization^4^.

Nevertheless, much of the volumetric brain study in the MRI era has not accounted for anthropomorphic normalization. Figures 2f and 2g show brain volume normalized by height-for-age and weight-for-height, respectively, which did not eliminate the sex-based differences in volume. Muscle mass content, greater in males, has been correlated with larger brain volumes although not with higher cognitive capability^30^. However, the ratio of brain to CSF volume demonstrated no significant sex differences at any age, without requiring anthropomorphic normalization (Figure 2h).

### Hemispheric Lateralization Findings

Figure 3a and 3b show that there was no lateralizing difference in size between right and left hemispheres for males and females, nor for cerebellum, frontal, parietal, or occipital lobes (Supplemental Figure 3). Figure 3c and 3d show that the left temporal lobe was significantly larger than the right for both males and females, which has been controversial^31^. In addition, the hippocampi were significantly larger on the right than the left side for both sexes (Figure 3e and 3f). Previous studies such as Lynch et al (2019) have also found that the right hippocampus is larger than the left^32^. Age-adjusted hippocampal and temporal lobe volume assessment may be of value in diagnosing and treating medically refractive epilepsy in childhood^31,33^.

### Correlation Between Cognition and Volume

Cognitive scores showed a small but significant correlation with brain volume in the four years leading up to the peak in volume (Figure 4). The Mental Development Index (MDI) scores for infants from birth to age 3 were not significantly predicted by brain volume (Figure 4b), but the Weschler Abbreviated Scale of Intelligence (WASI) scores for ages 6-18 years were significantly correlated with brain volume z-scores (Figure 4a), as described previously^34^. The correlation between cognitive score and brain volume was significant for raw brain volume, age-adjusted brain volume z-score, and weight-for-height normalized volumes in the years immediately preceding the brain volume peak (Figure 4c-4e). This correlation was not maintained when separating into smaller cohorts by sex (Supplemental Figure 4).

### Growth Curves and Centiles

In 1987, Roche et al created head circumference growth curves from studying 888 healthy US children^35^, and such normative head circumference curves from US^36^ and World Health Organization^27,37^ cohorts are now in routine clinical practice as indirect metrics of brain growth. Figure 4 illustrates the analogous GAMLSS pediatric brain volume growth curves for males and females (Figure 5a and 5b), with early brain volume growth and CSF volume insets included, as well as the brain/CSF ratio (Figure 5c and 5d) for a more comprehensive presentation of childhood normative brain growth suitable for clinical use.

**Figure 5.**
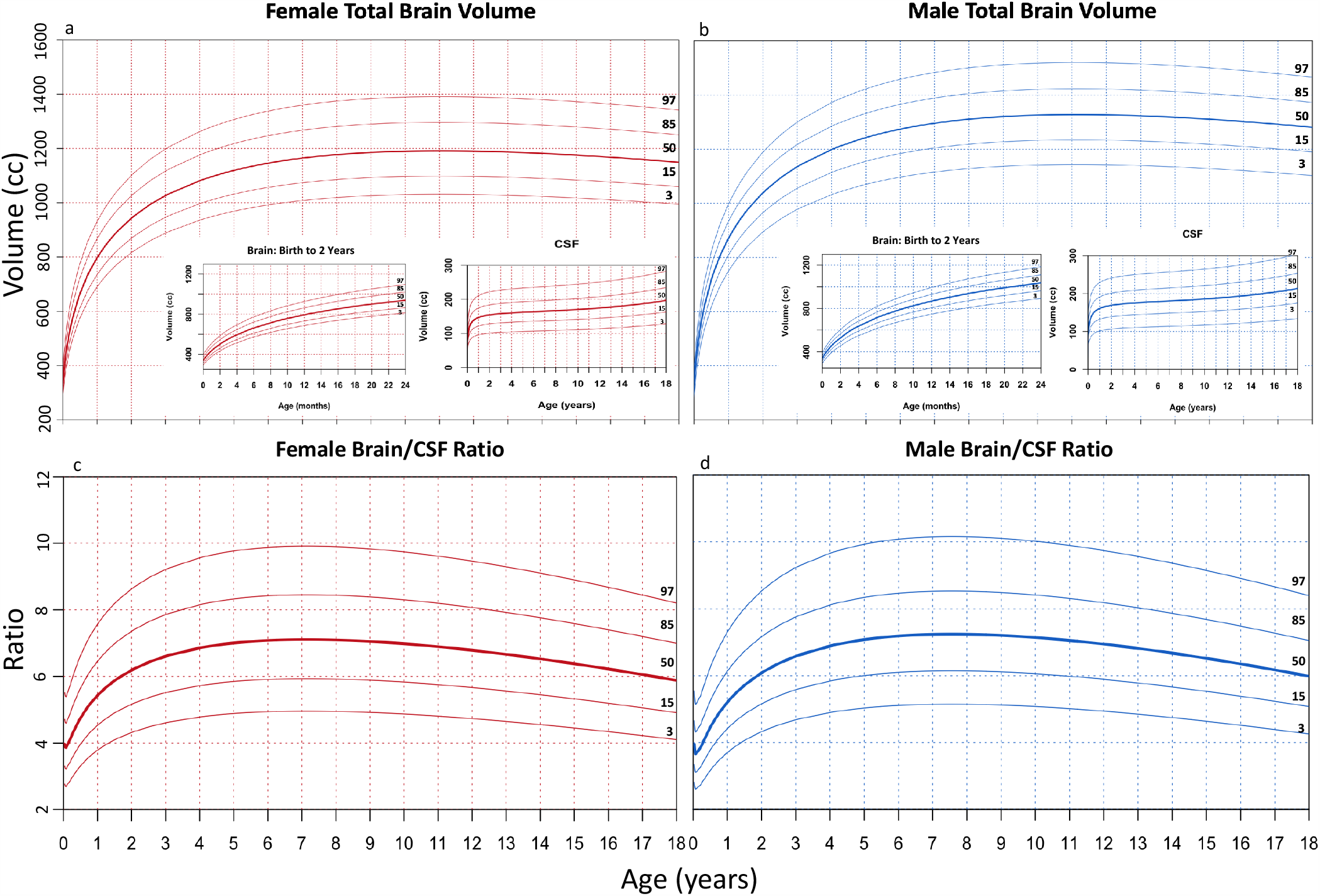
Standard Brain Volume Growth Curves. To provide a standard tool for researchers and clinicians, normal curves resembling head circumference growth curves were fit using GAMLSS to show the 3^rd^, 15^th^, 50^th^, 85^th^, and 97^th^ percentiles of normal brain volume growth for **a)** females and **b)** males, as well as the ratio of brain to CSF for **c)** females and **d)** males. The growth curves were modeled using a Box-Cox power exponential distribution and smoothed using fractional polynomials. Both male and female brain volume plots include insets for brain growth from birth to age 2, as well as CSF accumulation from birth to age 18, in order to provide a comprehensive picture of brain growth dynamics in the pediatric age range.

## Discussion

Brain volume measurement became a field of study of biological determinism pioneered by Samuel Morton in the mid-1800s^4,38^. Morton filled over 1000 cranial vaults with mustard seed and lead shot to determine brain volume, which he then compared between races and genders. A century and a half later Gould used Morton as a case study in scientific bias^4^. Decades after the publishing of these analyses in *The Mismeasure of Man*, arguments over biases and flaws continue in the assessment of the volume of the normal human brain^3^.

The sexual dysmorphism found in this present study corroborates previous studies looking at different age groups. Giedd et al (1999), Nopoulos et al (2000), Lenroot et al (2007), and Coupe et al (2017) all found similar differences in brain components, with larger total brain volumes in males^9,13,39,40^. We found that normalizing by body measurements did not account for sex differences; interestingly Coupe et al (2017) used proportionalization by total intracranial volume (TIV) and failed to find an overall sex difference^13^, a finding consistent with our discovery of a universal age-dependent ratio between brain and CSF volumes.

Another area of great interest in brain volume research is the growth trajectory of components of the brain. The head size increases steadily until at least age 18^35^. In the MRI era, however, many studies have shown that total brain volume reaches its peak in earlier adolescence. Without including younger children during the early years of rapid brain growth, the location of this volume peak can be somewhat uncertain. Giedd et al (1999) and Hedman et al (2012) showed that brain volume peaks between 12 and 14, but lacked subjects below age four when the majority of brain growth takes place. Lenroot et al (2007) found peaks at age 10.5 years for females and 14.5 for males, but lacked subjects below the age of three years^40^. Incorporating more children during the rapid early growth of the first few years of life our analysis placed the peak at 10-12 years of age.

It is interesting to note that Hedman et al (2012) found an early adolescent peak, but also concluded that their meta-analysis highlighted a possible later resurgence of growth with a secondary growth peak in adulthood^12^. These findings highlight the need for additional normal growth data throughout the life span. Such lifespan data would gain greater value by incorporating fetal through post-natal volume development, much in the manner of Holland et al (2014)^41^.

As with total brain volume, we found grey matter to peak in early adolescence and then slowly decrease. Our results showed that grey matter peaked in the seventh year of life for both males and females. Sowell et al (2003) also found that grey matter decreased after the seventh year of life, but they did not include any subjects under the age of seven^42^. While grey matter peaks in adolescence and starts to decline thereafter, the white matter and CSF components do not reach a peak during the pediatric age range. Sowell and others found that white matter reaches a peak near the age of 40 while CSF accumulation never ceases^28,42^. It is appreciated that myelination continues through the second decade of life contributing to white matter growth^43,44^. Grey matter, however, undergoes synaptic elimination through pruning, which may account for the peak seen in adolescence^45^. It is after the grey matter peak at seven years of age that we found a significant correlation between brain volume and cognitive scoring. This correlation continued until the total brain volume peaked near age 12. These findings highlight this critical period of pruning and myelination before overall brain volume begins to decline in late childhood.

In previous work, our group examined candidate growth models for brain^8^. One of the study limitations of that work was the small sample size number. In this present study, we have exhaustively incorporated what is, to our knowledge, the largest rigorously selected dataset of normal healthy children’s normal MRI scans. Constructing percentiled growth curves for clinical use, as opposed to model fitting for biology, requires accurate nonlinear curve fitting without respect to representing the underlying biology^35^. Figure 5 is, to our knowledge, the most definitive set of percentiled normal growth curves for brain for clinical use, along with percentiled curves for the brain/CSF ratio. Because it is currently unknown how to best incorporate these growth curves in clinical practice, we recommend using them within research protocols until best practices can be determined. We archive a high-quality growth chart in Supplementary Material to facilitate reproduction.

Our study has limitations. While the growth curves developed in this study provide a standard representation of the United States pediatric population, they can only act as a reference and not as a standard for other geographic regions. Other growth curves specific to particular regions and peoples should be developed using healthy cohorts derived from those regions in order to provide appropriately representative standards representing the diversity and characteristics of non-US populations.

## Conclusions

Our findings demonstrate that for a broad spectrum of human disease affecting neurocognitive development and brain growth – ranging from hydrocephalus to neonatal sepsis, intraventricular hemorrhage of prematurity, and childhood malnutrition – measuring brain growth and CSF with respect to normative values is now feasible. The apparent universal nature of the age-dependent brain/CSF ratio, regardless of sex or body size, suggests that the role of this ratio warrants further investigation. The finding that the brain/CSF ratio is tightly regulated opens up novel ways to characterize conditions affecting the childhood brain.

## Data Availability

The volumes and all of the data used to create the SSANOVA and GAMLSS growth curves presented here are provided in a spread sheet supplied in the online supplemental material (Supplemental Extended Data). The code needed to reproduce the figures in this study, as well as plot new volumes on the existing growth curves or calculate z-scores, is uploaded on the Github site https://github.com/Schiff-Lab/Brain-Growth

## Acknowledgements

We are grateful to T. Sauer and S. Sinnar for helpful discussion, and to Y. Wang and J. Chai for technical help in compiling data.

## Author Contributions

Conception and design: SJS, MRP. Acquisition of data: MRP, VC. Analysis and interpretation of data: MRP, VC, VM, JNP, PS, AK, BCW, SJS. Drafting the article: MRP, SJS. Critically revising the article: all authors. Reviewed and approved submitted version of manuscript: all authors.

## Supplemental Information

This work has previously been presented at the 2019 49th Annual Meeting of the AANS/CNS Section on Pediatric Neurological Surgery.

## Data Availability

The volumes and all of the data used to create the SSANOVA and GAMLSS growth curves presented here are provided in a spread sheet supplied in the online supplemental material (Supplemental Extended Data).

## Code Availability

The code needed to reproduce the figures in this study, as well as plot new volumes on the existing growth curves or calculate z-scores, is uploaded on the Github site https://github.com/Schiff-Lab/Brain-Growth

## Expanded Data

High Quality Reproduction of Figure 5: Normal Human Brain Growth and CSF Supplemental Excel File: Supplemental_Master_File.xlsm

## Supplemental Material

### Supplemental Methods

#### MRI Technical Details

Dual contrast T2-weighted (Fast/Turbo spin echo-ETL/Turbo factor 8, slice thickness 3 mm, oblique axial acquisition, TR 3300 ms, TEl 83 ms, TE2 165 ms, refocusing pulse 180 degrees, field of view and matrix AP: 256 & LR: 192 mm) scans were acquired for infants and Tl-weighted (3D RF-spoiled gradient recalled echo sequence, slice thickness 1 mm to 1.5 mm, sagittal acquisition, TR 22-25 MS, TE 10-11 ms, excitation pulse 30 degrees, whole head field of view, AP 256 matrix with LR for 1 mm isotropic) scans were acquired for adolescents. Each scan in the database was then corrected for intensity non-uniformities using the N3-algorithm in tandem with scanner-specific models of B0 inhomogeneity from each site^11^.

#### Smoothing Spline analysis of variance (SSANOVA)

Smoothing Spline analysis of variance (SSANOVA) is a semi-parametric method that models data generated from a smooth function *f*(*x*) by assuming that *f* is a function in a Reproducible Kernel Hilbert Space (RKHS) of the form ℋ = ℋ_0_ + ℋ_1_ ^1^. The set of functions 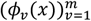, which spans the finite dimensional subspace ℋ_0_ and ℋ_1_, is a RKHS induced by a given kernel function *k*. Therefore, *f* has a semi-parametric form given by

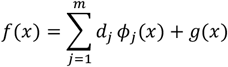

for coefficients *d*_*j*_, where functions *ϕ*_*j*_ have a parametric form and *g ∈* ℋ_1_ which is defined by:

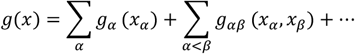

where *g*_*α*_ and *g*_*αβ*_ satisfy the standard ANOVA side conditions.

The SS-ANOVA estimate of *f* given data (*x*_*i*_, *y*_*i*_), *i* = 1, …, *n*, is given by the solution of a penalized problem,

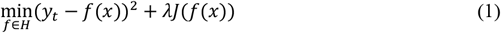

where the first term discourages the lack of fit of *f* and the second term penalizes the complexity of *f* with smoothing parameter *λ* controlling the trade-off between the two.

Following the representor theorem of Kimeldorf et al (1970)^6^ and the assumption of Gaussian data, the minimizer of the problem in equation (1) has a finite representation of the form:

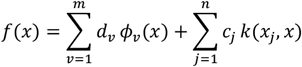

for coefficients *c*_*i*_ and *d*_*v*_.

Letting *Y* be the matrix of observations of size *N* × 1, where *N* includes all observations, including repeated measurements for different subjects; *S* is a *N* × *m* matrix where *m* represents the number of unpenalized terms in the model; and *Q* is a *N* × *N* matrix which accounts for all penalized terms in the model; estimation reduces to:

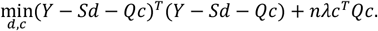

Here *S* is the matrix described above with the *iv*^*th*^ entry being *ϕ*_*v*_(*x*_*i*_) and *Q* is the penalized matrix with the *ij*^*th*^ entry being *k*(*x*_*i*_, *x*_*j*_)^3^. We use Generalized Approximate Cross-Validation (GACV), an approximation to the leave-one-out estimate of the comparative Kullback-Leibler distance between the empirical data 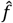 and the unknown true *f* to select regularization parameter *λ* and *θ*. This is dependent on the data available.

We were interested in visualizing and statistically analyzing the longitudinal differences between hemispheres, and therefore incorporated hemisphere as a main factor rather than a nested factor. Furthermore, some subjects only had one data point collected, while others had two or three. This imbalance and the low levels of repetition lead to matrix rank-deficiency in modeling hemisphere as a nested variable, rendering the nested approach mathematically inappropriate for this dataset.

#### Generalized Additive Models for Location, Scale, and Shape (GAMLSS)

The smooth growth curves used to fit the volumes and other growth metrics included in this study were developed using the Generalized Additive Models for Location, Scale, and Shape software implemented in R^10^. The Box-Cox power exponential (BCPE) distribution, which was chosen by the World Health Organization (WHO) for their standard growth curves, was used to model the volumes in this study^9^. This distribution models the median for a nonparametric assessment, and appropriately accounts for kurtosis and skewness within the data. The growth curves were fitted using the default RS algorithm and were smoothed using fractional polynomials of the third order^10^.

#### Segmentation

The neonates and infants were assessed using the Developing Human Connectome Project (dHCP) pipeline, which required T2 images and was run through a virtual Docker container to access a Linux computer system^8^. The dHCP pipeline relies on the Brain Extraction Tool (BET) algorithm for skull-stripping, followed by the N4 algorithm for intensity inhomogeneity correction and the Draw-EM algorithm for segmentation ^7,12,13^. The Draw-EM (Developing brain Region Annotation With Expectation-Maximization) algorithm consists of an adaptive Expectation-Maximization scheme relying on a spatial prior term and intensity model. The spatial prior term is established using labeled atlases that are registered to the image, whereas the intensity model is established using a Gaussian Mixture Model. The labeled atlases were obtained through manual editing and later updated by Markopoulos et al^4,7^.

The older subjects were assessed using the Computational Anatomy Toolbox 12 (CAT12) within the Statistical Parametric Mapping (SPM) platform using Matlab 2019b, which relies on T1 images^2^. The CAT12 algorithm relies on voxel-based morphometry for segmentation and label-based morphometry for region-of-interest (ROI) classification. Each scan was preprocessed using a modified ICBM Tissue Probabilistic Atlas tissue probability map along with an ICBM space template affine registration. Skull-stripping was accomplished using adaptive probability region-growing (APRG) algorithm followed by surface-based optimization. The MNI152 Dartel Template was used for Dartel registration, and finally the Hammers atlas was used for ROI analysis^5^. Each of the resulting scan sets was manually curated to ensure that appropriate skull-stripping and segmentation was accomplished. Upon establishing the volumes determined by each segmentation procedure, the accompanying atlases were used to compile volumes for the desired regions from smaller sections of the brain^4,5,7^.

Upon establishing the volumes determined by each segmentation procedure, the accompanying atlases were used to compile volumes for the desired regions from smaller sections of the brain^4,5,7^.

### Supplemental Figures

**Supplemental Figure 1.**
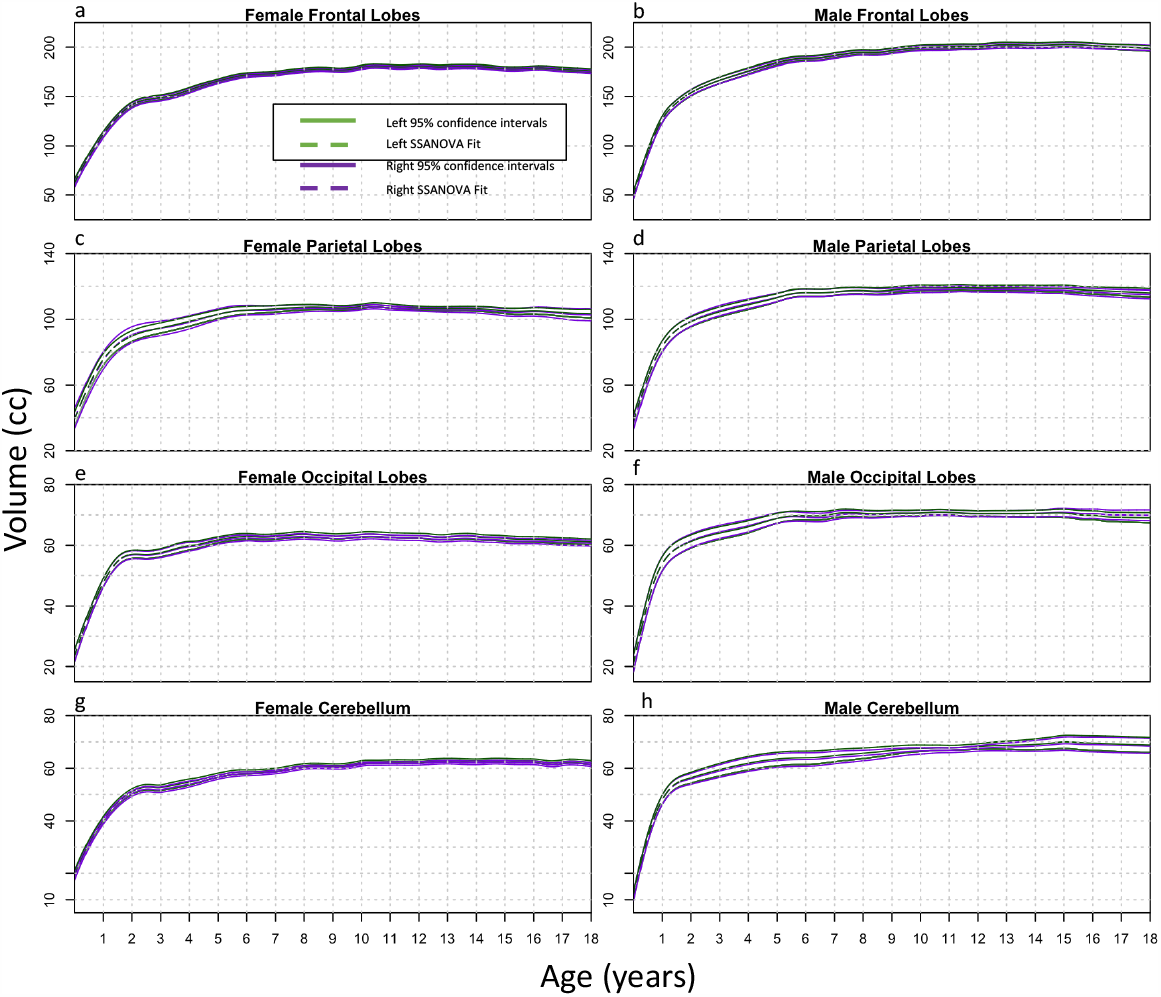
Brain Lobe and Cerebellum Hemispheric Differences. SSANOVA models with random effects were fit to **(a**,**b)** frontal lobes (cc), **(c**,**d)** parietal lobes (cc), **(e**,**f)** occipital lobes (cc), **(g**,**h)** and cerebella (cc). Female regional growth curves from birth to 18-years-old are shown in the left column, and the corresponding male regional curves are shown in the right column. For each plot, the left side is shown in green and the right side in purple. The dashed line represents the fit, with the two solid lines on either side showing the Bayesian 95% confidence intervals. The upper and lower intervals for the right and left sides overlap throughout the entire time frame for each region, and therefore there are no statistically significant differences.

**Supplemental Figure 2.**
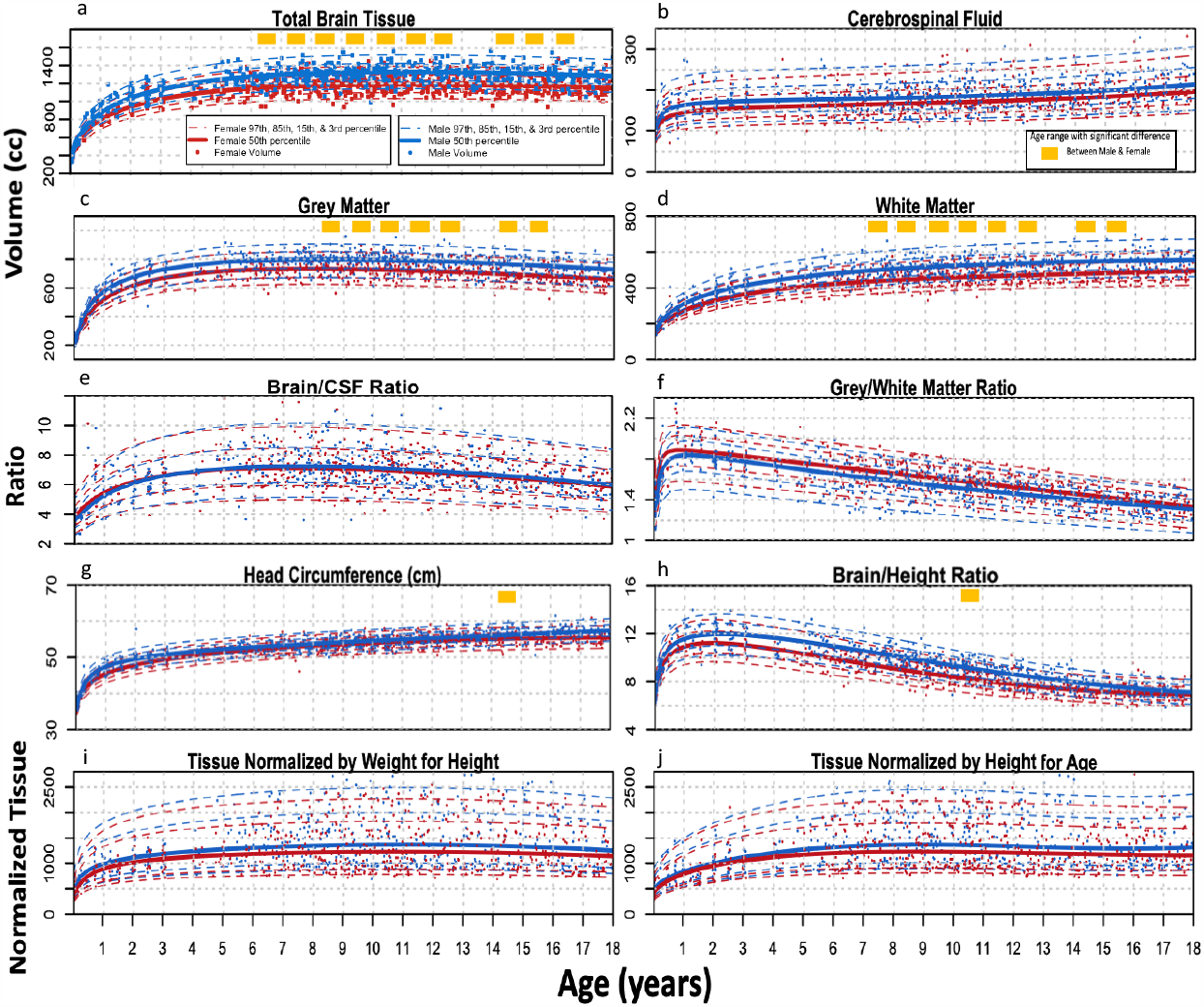
Brain Component Standard Growth Curves. Growth curves from birth to 18-years-old developed with Generalized Additive Models for Location, Shape, and Scale are shown for males in blue and females in red, with the solid median accompanied by dotted one and two standard deviations above and below. Each growth curve was modeled using a Box-Cox power exponential distribution and smoothed using fractional polynomials. The yellow boxes represent Bonferroni corrected significant differences (p<0.00006) between males and females for each year, obtained using the Mann-Whitney test. This univariate testing was applied to obtain conservative estimates of the differences in the data for each year of life. The standard curves plotted include **a)** total brain volume (cc), **b)** CSF, **c)** grey matter, **d)**white matter, **e)** the ratio of total brain volume to CSF, **f)** the ratio of grey to white matter, **g)** head circumference (cm), **h)** height normalized brain volume, **i)** weight for height normalized brain volume, and **j)** height for age normalized brain volume.

**Supplemental Figure 3.**
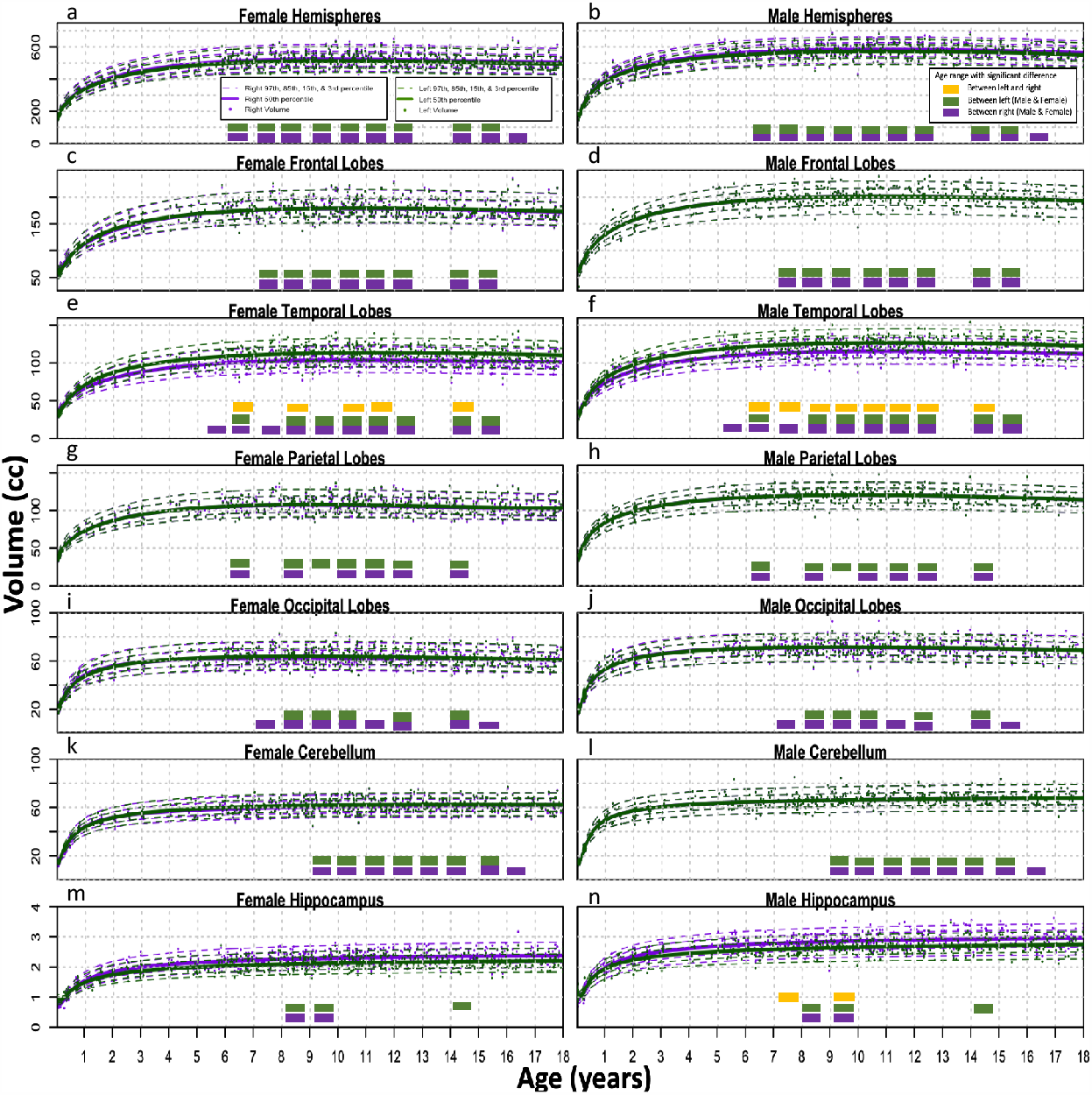
Brain Region Standard Growth Curves. Female regional growth curves from birth to 18-years-old are shown in the left column, and the corresponding male regional curves are shown in the right column, all developed using Generalized Additive Models for Location, Shape, and Scale. Each growth curve was modeled using a Box-Cox power exponential distribution and smoothed using fractional polynomials. Left regions are shown in green, and right regions are shown in purple, with the solid median accompanied by dotted one and two standard deviations above and below. The yellow boxes represent Bonferroni corrected significant differences (p<0.00006) between left and right components in that year, obtained using the Mann-Whitney test. The green boxes represent significant differences between the left region for males and females in that year, while the purple boxes represent significant differences between the right region for males and females in that year. The standard curves plotted include **(a**,**b)** hemispheres (cc), **(c**,**d)** frontal lobes (cc), **(e**,**f)** temporal lobes (cc), **(g**,**h)** parietal lobes (cc),**(i**,**j)** occipital lobes (cc), **(k**,**l)** cerebella (cc), and **(m**,**n)** hippocampi (cc).

**Supplemental Figure 4.**
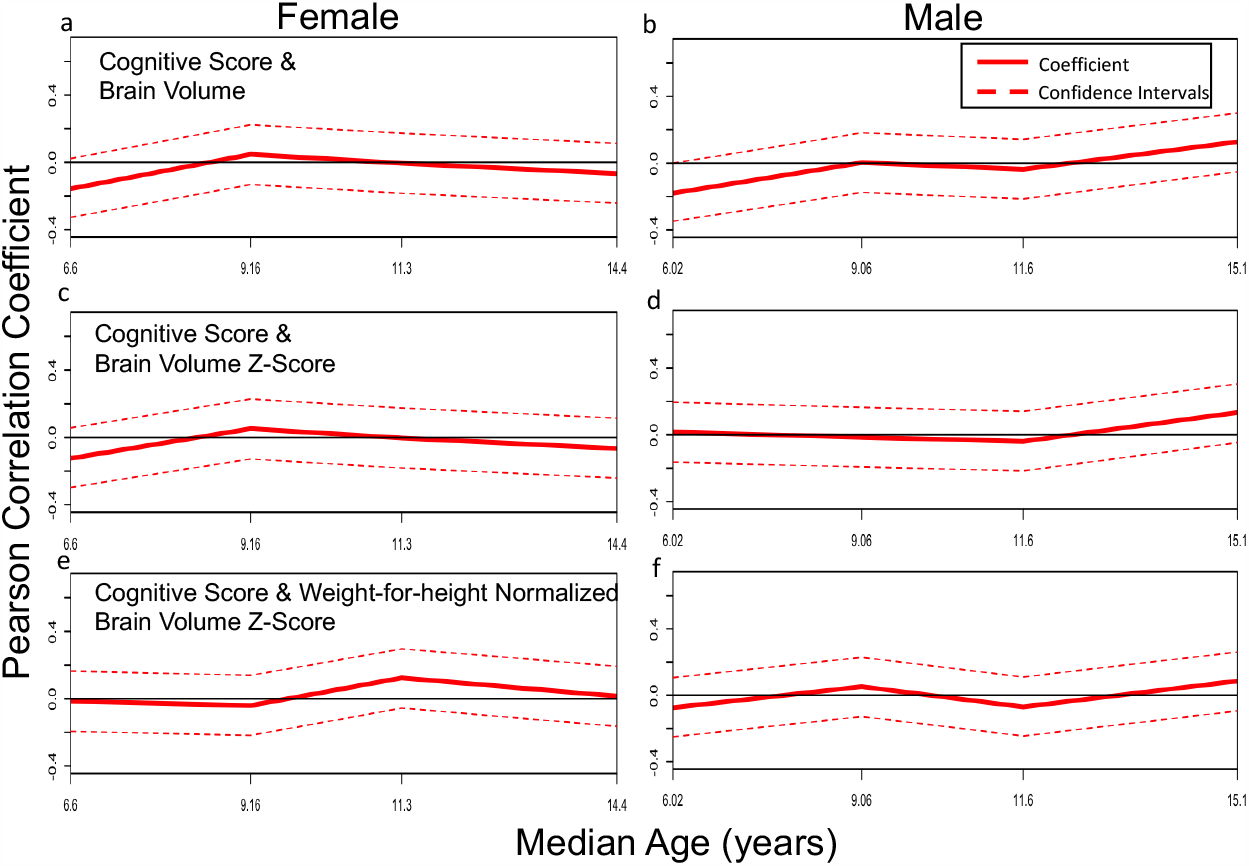
Cognitive Score Correlations by Sex. Age-dependent correlations were calculated with a subpopulation window-size of 120 and step-size of 20 subjects to show the Pearson Correlation and confidence intervals between cognitive score and **(a**,**b)** raw brain volume, **(c**,**d)** brain volume z-score, and **(e**,**f)** weight-for-height normalized brain volume z-score at different ages for both females and males.

